# Resolution of systemic inflammation in psoriasis following herring roe oil treatment: a *post hoc* analysis on inflammatory biomarkers in non-severe psoriatic patients

**DOI:** 10.64898/2026.04.20.26350934

**Authors:** Thomas Ringheim-Bakka, Runhild Gammelsaeter, Kåre Steinar Tveit

## Abstract

**Background:** Psoriasis is a chronic immune-mediated inflammatory disease (IMID) with systemic involvement. In mild-to-moderate disease, circulating cytokines may inadequately capture systemic inflammatory burden. Composite haematological indices derived from complete blood counts, such as the systemic immune–inflammation index (SII) and systemic inflammation response index (SIRI), have emerged as sensitive prognostic markers of systemic inflammation, including in psoriasis. This exploratory *post hoc* analysis investigated the effects of orally administered herring roe oil (HRO), a phospholipid-rich marine oil, on systemic inflammation in patients with mild-to-moderate psoriasis utilizing these biomarkers.

**Methods:** Data were analysed from a randomized, double-blind, placebo-controlled 26-week clinical study which investigated HRO supplementation in patients (N = 64) with mild-to-moderate psoriasis (NCT03359577). SII, SIRI, neutrophil-to-lymphocyte ratio (NLR), platelet-to-lymphocyte ratio (PLR), and monocyte-to-lymphocyte ratio (MLR) were calculated at baseline, week 12, and week 26 for patients where baseline complete blood counts (CBCs) were available (n = 60). Patients missing baseline CBCs were excluded from the analysis. Continuous changes were assessed using ANCOVA with baseline adjustment. Categorical responder analyses were performed with 25% and 30% reduction thresholds and stratification by baseline biomarker medians were performed to evaluate treatment responses and impact of baseline inflammation.

**Results:** Compared with placebo, HRO treatment resulted in significant mean reductions in SII, SIRI, and PLR at week 26, with supportive trends and responder effects observed as early as week 12 compared to placebo. Patients with elevated baseline inflammatory indices showed the greatest reductions in systemic inflammation. Stratification by baseline SII further revealed enhanced clinical benefit, with statistically significant PASI50 response rates in the HRO arm at week 26 among patients with lower baseline SII.

**Conclusion:** HRO supplementation was associated with a time□dependent reduction in systemic inflammatory biomarkers in mild□to□moderate psoriasis patients. These findings support the utility of composite inflammatory indices for monitoring systemic inflammation and suggest that baseline SII may have utility in predicting treatment response and may be a useful tool for stratification in clinical trials in mild to moderate psoriasis patients. These results could also suggest platform-potential of HRO for resolution□oriented interventions across several inflammatory conditions.

## 1 Introduction and background

Psoriasis is a chronic, immune-mediated inflammatory disease (IMID) affecting the skin and is most known and recognized by its hallmark raised, inflamed and scaly lesions. The prevalence of psoriasis, where it affects roughly 2-4% of the Western population, makes it a widespread disease with millions of patients across the globe, and it made up close to 7% of all new IMID cases in 2019 (1,2). Out of all patients with a confirmed psoriasis diagnosis, around 80% of cases in the US are reported as mild disease, who are usually not eligible for systemic or biologic treatments and have a significant unmet need for new treatment options (3). Although psoriasis is characterized as a skin disease, it is increasingly recognized to have systemic impact with inflammatory processes extending beyond the skin niche (4). Consistent with systemic immune-driven inflammation, patients with psoriasis have increased risk for a range of comorbidities, including cardiovascular disease, gastrointestinal disease, and psoriatic arthritis (5).

The systemic impact of psoriasis is evident by elevated levels of circulating inflammatory cytokines such as TNF-alpha, IL-17 and -23. However, in non-severe psoriasis populations it may be challenging to assess systemic inflammation, as the cytokine levels though elevated in the skin lesions may not always be reliably reflected in circulating cytokine measurements. It was recently shown that peripheral blood mononuclear cells from mild-to-moderate psoriasis patients and healthy controls were found to express IL-17A and IL-17F at low and comparable levels (6). Considering the inconsistent impact on circulating inflammatory cytokines in non-severe psoriatic disease populations, it may be challenging to assess the systemic impact of their inflammatory disease using circulating cytokine levels.

Alternative assessments of systemic inflammation to measuring circulating cytokines in milder disease populations are systemic composite inflammatory biomarkers, such as systemic immune inflammation index (SII) and systemic inflammation response index (SIRI). These biomarkers are emerging as sensitive, low-cost, and non-invasive indices calculated from complete blood count (CBC) values for assessing systemic inflammation in peripheral blood, where SII is calculated by neutrophils x platelets / lymphocytes and SIRI by neutrophils x monocytes / lymphocytes (7). Although originally being developed for predicting prognosis and overall survival in oncology, SII and SIRI have now been explored in many other indications, including cardiovascular disease and diabetic depression (8–12). Furthermore, emerging evidence are suggesting that elevated SII and SIRI is associated with risk of multiple autoimmune and IMID conditions and are also associated with disease activity in several of these conditions (13–15). Both elevated SII and SIRI have been found to be associated with risk of psoriasis, psoriasis disease activation, and psoriatic disease activity (16–20). Systemic biological treatment for psoriasis has furthermore been observed to lead to reduction in SII over time (21). In addition to being a prognostic biomarker, SII may also be a promising predictor for treatment trajectories, as elevated baseline SII has been associated with poor treatment outcomes and treatment switches in patients with moderate-to-severe psoriasis receiving biologic treatments (22).

Considering the growing body of evidence linking these systemic inflammatory indices to psoriasis disease risk, disease activity, and treatment outcomes in a plethora of indications with an inflammatory component - including psoriasis - it was of interest to assess impact of SII and SIRI in an existing clinical dataset where herring roe oil (HRO) had been evaluated for its potential in treating non-severe psoriasis. The randomized, double-blind, placebo-controlled 26-week clinical study which investigated HRO supplementation versus placebo (NCT03359577) was conducted in patients with mild-to-moderate psoriasis (N = 64). The primary outcome of the trial showed that patients in the HRO arm experienced a statistically significant greater mean reduction in psoriasis area severity index (PASI) compared to placebo at 26 weeks (23). A previous *post hoc* cytokine analysis of data from this study also showed that HRO had systemic effects on immune cells, where reduction in CCL2 levels and an increase in IFN-γR1 was observed over time (24). With the significant improvement in skin symptoms and indications of impact of HRO on immune cells and cytokine network, it was hypothesised that an impact on SII and SIRI could potentially be observed.

The interventional treatment in the HRO study was a marine phospholipid (PL)-rich oil extracted from herring (*Clupea harengus*) roe enriched in marine polyunsaturated fatty acids (PUFAs). The long-chain PUFAs (LC-PUFAs) in HRO are primarily docosahexaenoic acid (DHA) and eicosapentaenoic acid (EPA) in a 3:1 ratio. The key differentiator of HRO compared to more traditional fish oils is the high content of marine phospholipid esters enriched in marine LC-PUFAs, where marine PLs have been implied to have superior bioavailability and be actively transported across the blood-brain barrier, as well as having anti-inflammatory effects (25–33). Specifically, it has previously been shown that HRO supplementation reduces peripheral inflammation and circulating neutrophils in a preclinical piglet model as well as improving peripheral blood n-6/n-3 ratios in healthy volunteers (34,35). HRO has also been observed in a recent publication and a preprint to promote biosynthesis of specialized pro-resolving mediators (SPMs) and limit IL-23 secretion in macrophages *in vitro* (36,37). All these observations are suggestive of a potential impact of HRO treatment on systemic inflammatory indices such as SII and SIRI.

Based on previous results from this trial and the prior knowledge of HRO, the aim of this exploratory *post hoc* analysis was to investigate the potential impact of HRO supplementation on SII and SIRI in mild-to-moderate psoriasis patients in a placebo-controlled clinical setting. It was also considered relevant to assess a potential impact of HRO treatment on the simpler composites neutrophil-to-lymphocyte ratio (NLR), platelet-to-lymphocyte ratio (PLR), and monocyte-to-lymphocyte ratio (MLR) as they incorporate similar cell types to SII and SIRI, and have also been associated with active psoriasis disease (38). Specifically, where NLR and PLR have been positively correlated with mean PASI scores, while MLR, together with NLR, PLR, and SII, has been associated with the presence of psoriasis. In addition to analysing the direct impact on the composite ratios through mean change analyses or categorical response threshold analyses, it was of interest to stratify patients based on baseline values for the different systemic inflammatory indices to assess if degree of inflammatory load at baseline impacted treatment trajectories both with regards to change in systemic inflammation as well as change in skin symptom severity and quality of life (QoL) impact. If so, inflammatory biomarker status at baseline could be a potential predictor of response to HRO treatment.

## 2 Materials and methods

### 2.1 Trial dataset details and population

This exploratory *post hoc* investigation on composite systemic inflammation biomarkers was performed on the dataset from the HRO study (clinicaltrials.gov identifier: NCT03359577). The HRO study was a double blinded, single-centre, randomized, and placebo-controlled 26-week trial conducted in mild-to-moderate psoriasis patients at the Department of Dermatology at Haukeland University Hospital in Norway. After the placebo-controlled period, a selection of patients elected to continue with an open-label extension (OLE) for 39 weeks where all were assigned to HRO treatment. The OLE period was not subject to biomarker analysis in this investigation due to the lack of placebo control. The present study adheres to the trial’s ethical approvals and consent procedures as reported previously (23).

Mild-to-moderate psoriasis patients (N = 64) were recruited and randomized (1:1) to receive either herring roe oil (HRO) or matching placebo capsules filled with medium chain triglycerides (MCT) for 26 weeks. Patients eligible for recruitment were adults (≥18 years) with chronic and active mild- to-moderate plaque psoriasis (PASI <10). Patients on systemic anti psoriatic therapy or immunosuppressive/immunomodulating treatments known to influence psoriasis were excluded from participation in the trial.

Each soft capsule of HRO contained 292 mg polyunsaturated fatty acids with a 1:3 ratio of eicosapentaenoic acid (EPA) and docosahexaenoic acid (DHA), leading to a total dose of 1.9 g/day of DHA and 0.7 g/day of EPA. Approximately 35% of the fatty acids were bound in marine phospholipids with phosphatidylcholine being the main phospholipid. The recruited patients were administered 10 capsules per day, resulting in a total lipid dose of 5.9 g/day over the 26 weeks of treatment.

This *post hoc* composite biomarker investigation required CBC samples for the patients at the three visits (week 0, week 12, and week 26). Four patients (2 active and 2 placebo) were missing baseline CBC values and were thus excluded from the analysis, resulting in a total population of 60 patients with 1:1 distribution between HRO and placebo. The baseline characteristics of the 60 patients and their skin symptom and QoL development throughout 26 weeks of treatment is shown in Table 1 and Figure 1. Missing data after 12 weeks and 26 weeks of treatment for the 60 patients was consistent and remained below 10%. Because of the low level of missingness, no methods to address missing data were applied, and all analyses were conducted on data as observed without imputation.

**Table 1:**
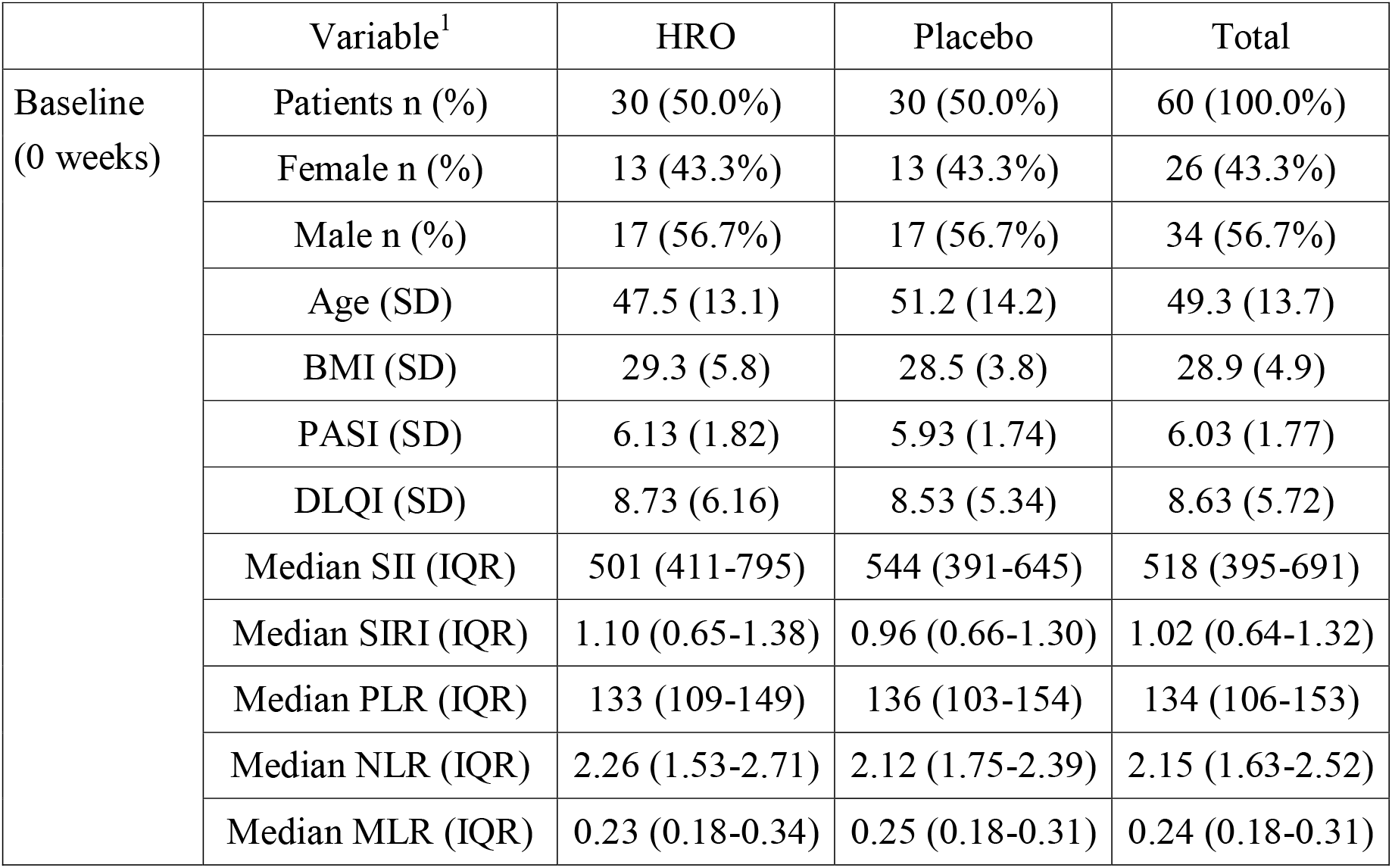
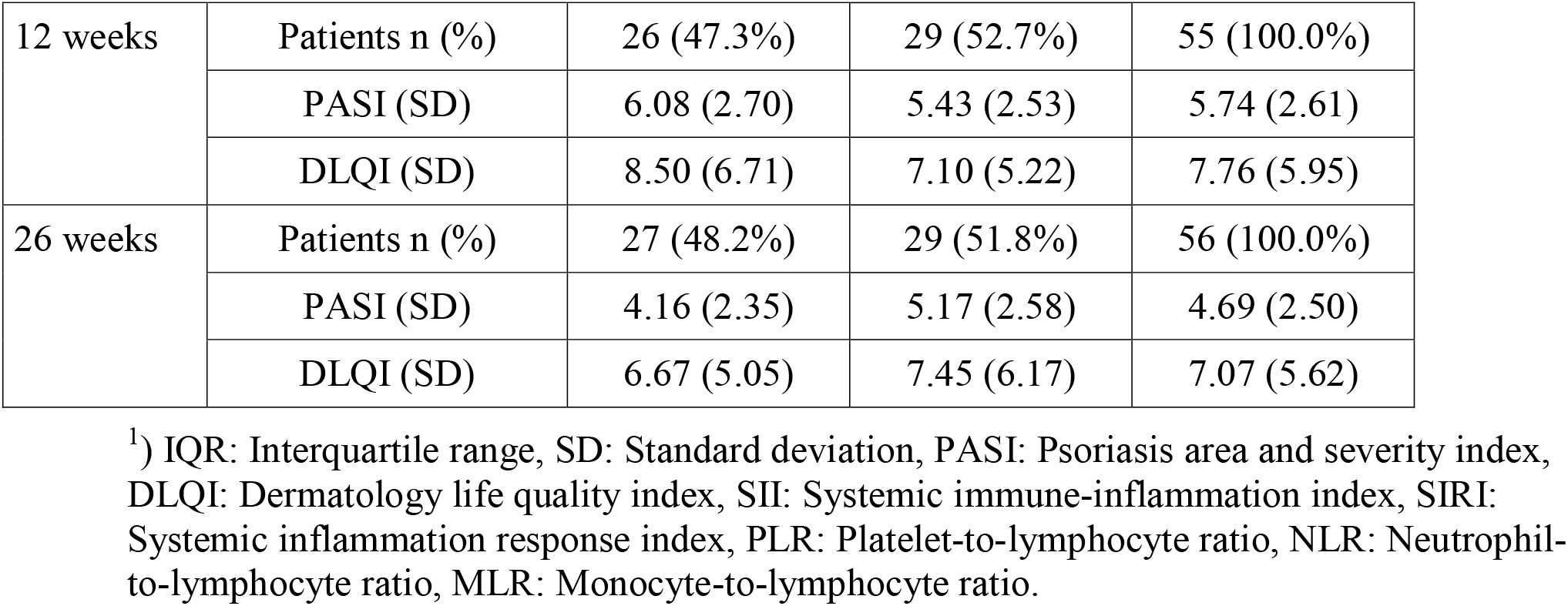
Baseline characteristics (n = 60) for patients eligible for composite inflammatory biomarker analysis and development of skin symptoms and patient QoL over 26 weeks of treatment.

**Figure 1:**
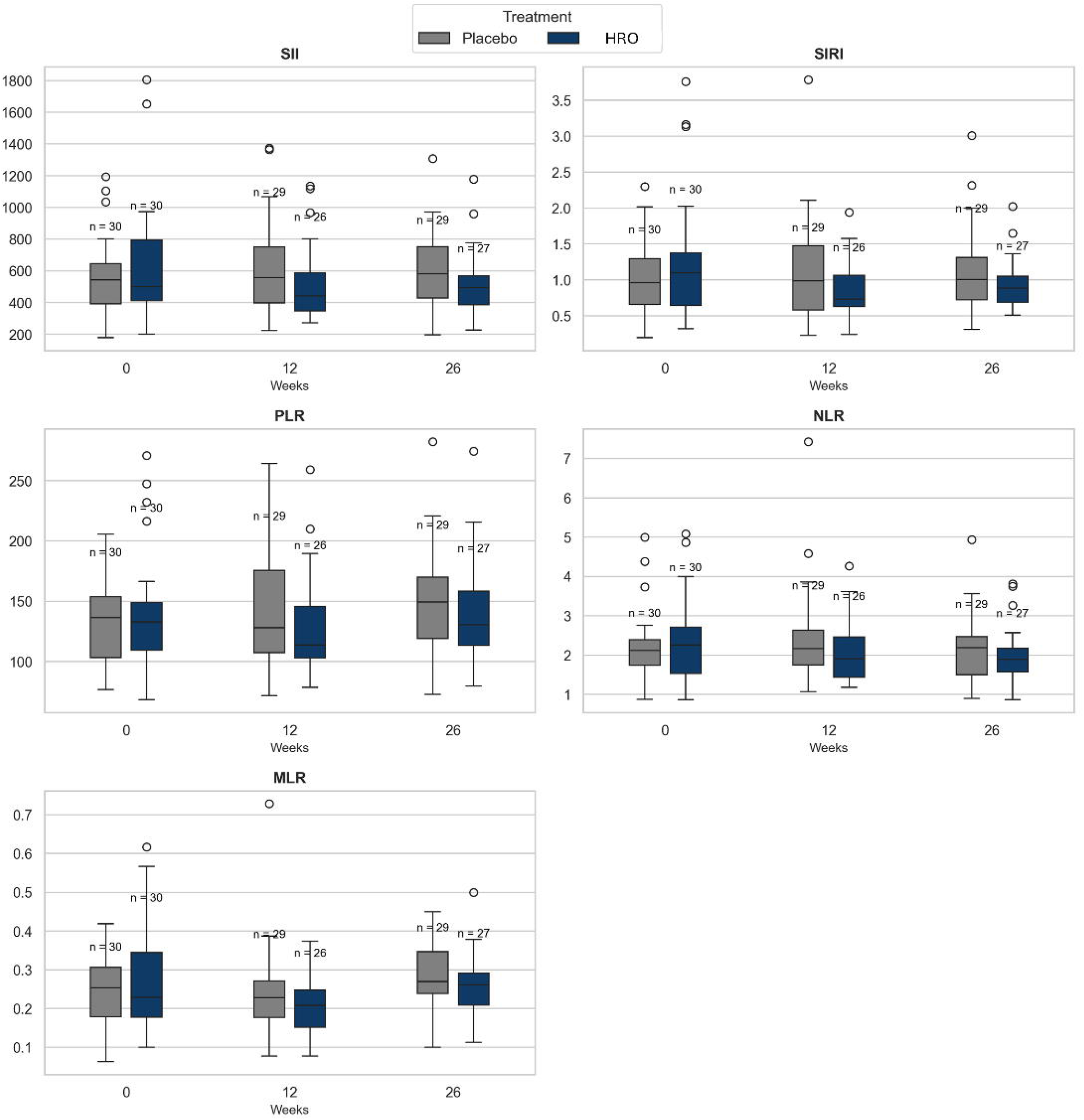
Multi-panel boxplot for SII, SIRI, PLR, NLR, and MLR in the HRO and placebo groups at baseline (week 0) and at week 12 and week 26.

### 2.2 Blood samples, clinical examinations, and biomarker analysis

Details on blood sampling and clinical examinations has been described by Tveit *et al*. (23), and composite biomarker values were calculated from existing CBC data.

The composite biomarkers evaluated were systemic immune-inflammation index (SII; neutrophils x platelets / lymphocytes), systemic inflammation response index (SIRI; neutrophils x monocytes / lymphocytes), neutrophil-to-lymphocyte ratio (NLR; neutrophils / lymphocytes), platelet-to-lymphocyte ratio (PLR; platelets / lymphocytes), and monocyte-to-lymphocyte ratio (MLR; monocytes / lymphocytes).

### 2.3 Patient stratification

The patient population was divided into multiple subgroups based on whether they were above or below the baseline median SII, SIRI, PLR, NLR, or MLR for all patients. The population median was used to objectively define the cutoff without influence from treatment arm-specific medians even if it skewed the distributions. The subgroups above and below the median baseline values were separately assessed for responder proportions compared to placebo for a range of categorical endpoints.

### 2.4 Statistical methods

Continuous change in composite inflammatory biomarkers at weeks 12 and 26 was analysed using analysis of covariance (ANCOVA) with the corresponding baseline value included as a covariate. Ordinary least squares estimation with classical standard errors was applied.

Categorical threshold analysis of 25% and 30% reduction of SII, SIRI, NLR, PLR, and MLR were analysed using the Pearson chi-square test; when any cell counts in the 2×2 contingency table were ≤5, Fisher’s exact test (two-sided) was used instead. The 25% threshold was defined based on observations showing mean changes of similar magnitudes in SII, NLR, PLR, and MLR over time for patients receiving biological psoriasis treatments (21,39), and the 30% threshold was included as a sensitivity analysis. Odds ratios (ORs) were calculated as the cross□product ad/bc when all four cells were non□zero; when any cell was zero, the Haldane–Anscombe 0.5 continuity correction was applied to all four cells. Ninety-five percent confidence intervals for categorical endpoints were calculated using the Wald (Woolf) method for Pearson chi-square analyses and the Baptista–Pike method when the Fisher’s exact test was applied. Statistical significance was defined as p□<□0.05 for all analyses.

All analyses were executed locally in Microsoft VS Code (v1.110.0, Windows 11) using Python 3.11. The following Python packages were used: pandas, numpy, statsmodels, patsy, scipy, matplotlib. Scripts will be made available upon reasonable request.

## 3 Results

### 3.1 HRO treatment reduces systemic inflammatory biomarkers

HRO treatment resulted in statistically significant reduction in several inflammatory indices compared to placebo at week 26 (Table S1). Significant decreases were observed for SII (ΔHRO vs ΔPBO: -100.32; 95% CI: -190.38 to -10.27; p = 0.0297), SIRI (ΔHRO vs ΔPBO: -0.26; 95% CI: - 0.48 to -0.03; p = 0.0263), and PLR (ΔHRO vs ΔPBO: -17.38; 95% CI: -33.24 to -1.52; p = 0.0323) in addition to a nonsignificant trend for MLR (ΔHRO vs ΔPBO: -0.03; 95% CI: -0.07 to 0.00; p = 0.0734). No significant reduction in the HRO group compared to placebo was observed for NLR (ΔHRO vs ΔPBO: -0.18; 95% CI: -0.46 to 0.10; p = 0.1988) at week 26. No significant reductions were observed in the HRO group at week 12 compared to placebo. However, trends towards reductions were noted for MLR (ΔHRO vs ΔPBO: -0.05; 95% CI: -0.10 to 0.00; p = 0.0717) and PLR (ΔHRO vs ΔPBO: -15.92; 95% CI: -34.04 to 2.20; p = 0.0838), indicating that onset of inflammation resolution may have already initiated after 12 weeks but failed to achieve significant reduction before week 26. Unadjusted changes in composite biomarker levels across the treatment period are shown in Table S2.

### 3.2 Inflammatory biomarker responder rates support continuous-scale reductions

Categorical responder analyses were performed with 25% reduction (SII25, SIRI25, NLR25, PLR25, and MLR25) as the key measure for the different inflammatory biomarkers. Results from these analyses are shown in Figure 2 and Table S3 together with results from the sensitivity analyses at 30% reduction below.

**Figure 2:**
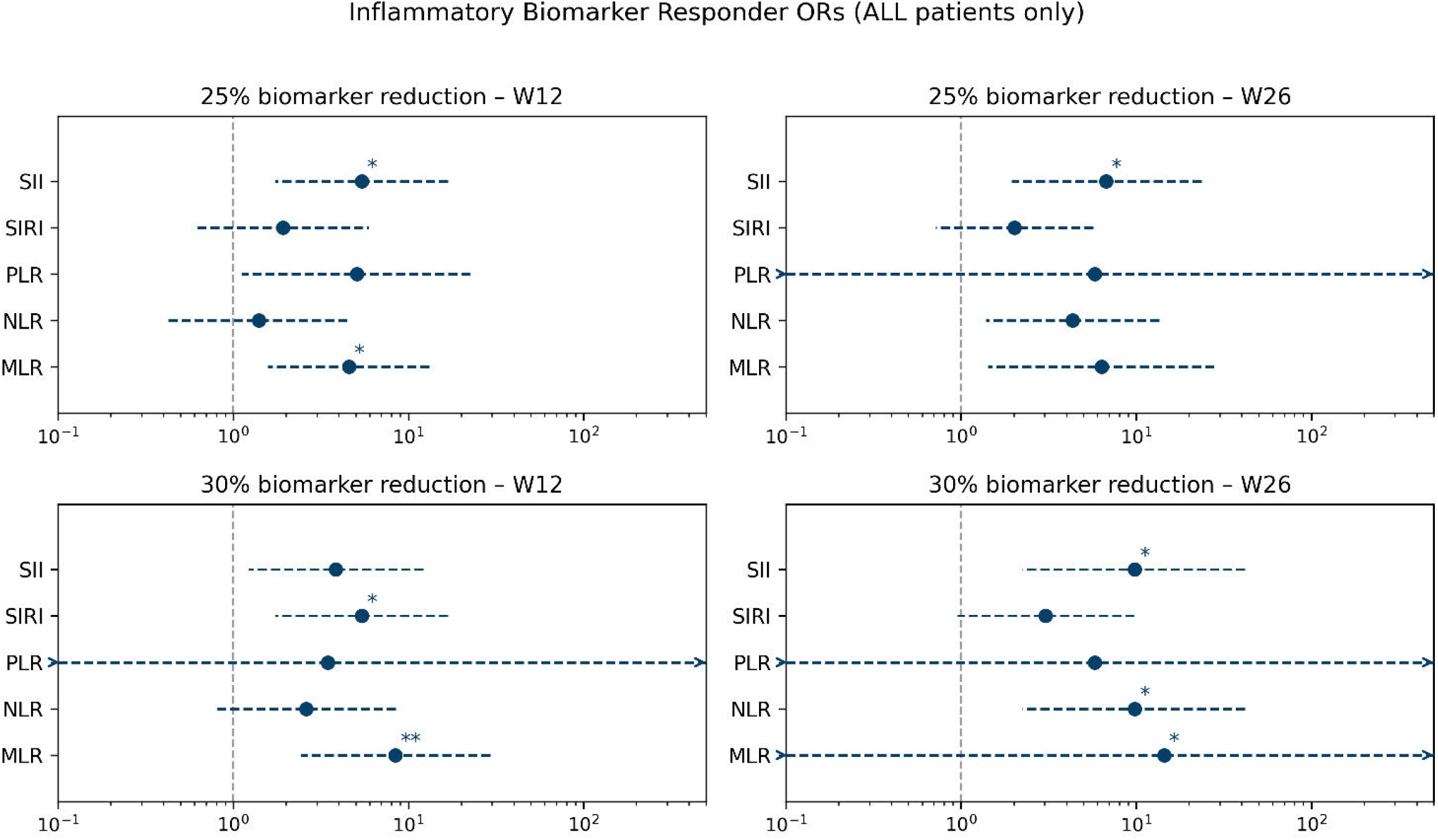
Forest plots showing responder proportions for 25% and 30% reduction of the composite biomarkers as the odds ratio (OR) between HRO and placebo at week 12 (W12) and week 26 (W26). ORs >1 is in favour of response in the HRO group. *) p < 0.05 and **) p < 0.01 according to the Fisher exact test. Whiskers denote 95% CIs and where exact CIs are unbounded, whiskers are truncated at the axis and arrows indicate that the CI extends beyond the plotted range.

The responder rates in the HRO group were found to be statistically significant compared to placebo already after 12 weeks for SII25 at 38% vs 10% (OR: 5.42; 95% CI: 1.74 to 16.81; p = 0.0245) and MLR25 at 42% vs 14% (OR: 4.58; 95% CI: 1.57 to 13.16; p = 0.0321). After week 26 the only statistically significant responder rate in the HRO group compared to placebo was found for SII25 at 33% vs 7% (OR: 6.75; 95% CI: 1.94 to 23.63; p = 0.0184). Though it should be noted that NLR25 had marginally significant responder rates at 33% vs 10% (OR: 4.33; 95% CI: 1.39 to 13.56; p = 0.0519).

Sensitivity analyses were then conducted using a stricter 30% threshold to assess whether the HRO treatment effect was sustained or diminished under more stringent responder criteria. Tightening the response criteria to 30% reduction gave statistically significant responder proportions in the HRO group compared to placebo at week 12 for SIRI30 at 38% vs 10% (OR: 5.42; 95% CI: 1.74 to 16.81; p = 0.0245) and MLR30 at 38% vs 7% (OR 8.44; 95% CI: 2.43 to 39.30; p = 0.0076). At week 26 statistically significant response rates in the HRO group compared to placebo were observed for SII30 and NLR30 both at 26% vs 3% (OR: 9.80; 95% CI: 2.23 to 41.87; p = 0.0227) and MLR30 at 19% vs 0% (OR: 14.42; 95% CI: 0.00 to inf.; p = 0.0211).

### 3.3 Baseline systemic inflammation stratified by biomarkers is associated with clinical treatment response

It was furthermore of interest to examine whether baseline inflammatory biomarker levels influenced the magnitude or direction of treatment responses. The above- and below-median subgroups were then evaluated based on responder proportions for 25% and 30% biomarker reduction, PASI50, and achievement of the minimal clinically important difference in QoL, defined as a ≥4-point reduction in DLQI (Table S4; Table S5). Including skin symptom assessments and QoL metrics was done to evaluate treatment trajectories on clinical symptoms in addition to the magnitude and direction of the biomarker responses.

Dividing the population in two based on the median SII revealed a statistically significant response rate in the HRO group compared to placebo for achieving PASI50 at week 26 in the group with baseline SII ≤518, with responder proportions at 44% vs 8% (OR: 9.33; 95% CI: 1.99 to 39.23; p = 0.0443). Statistically significant PASI50 responder proportions were not observed in the HRO group for the entire population, nor in any of the other subgroups divided based on the median SIRI, PLR, NLR or MLR. Nor were any significant results observed for reduction of DLQI in any of the subgroups.

Analysis of biomarker reduction in the subgroups with baseline biomarkers above the median resulted in >30 statistically significant responder proportions in the HRO arm compared to placebo at week 12 and week 26 (Figure 3). At week 12, all the statistically significant reductions in the HRO group compared to placebo were observed for SII, SIRI, and MLR with different magnitudes of reduction and across all subgroup definitions (i.e. above baseline median SII, SIRI, PLR, NLR, and MLR). At week 26, the statistically significant responder proportions were mainly observed for SII and NLR (9 of each), whereas none were observed for PLR.

**Figure 3:**
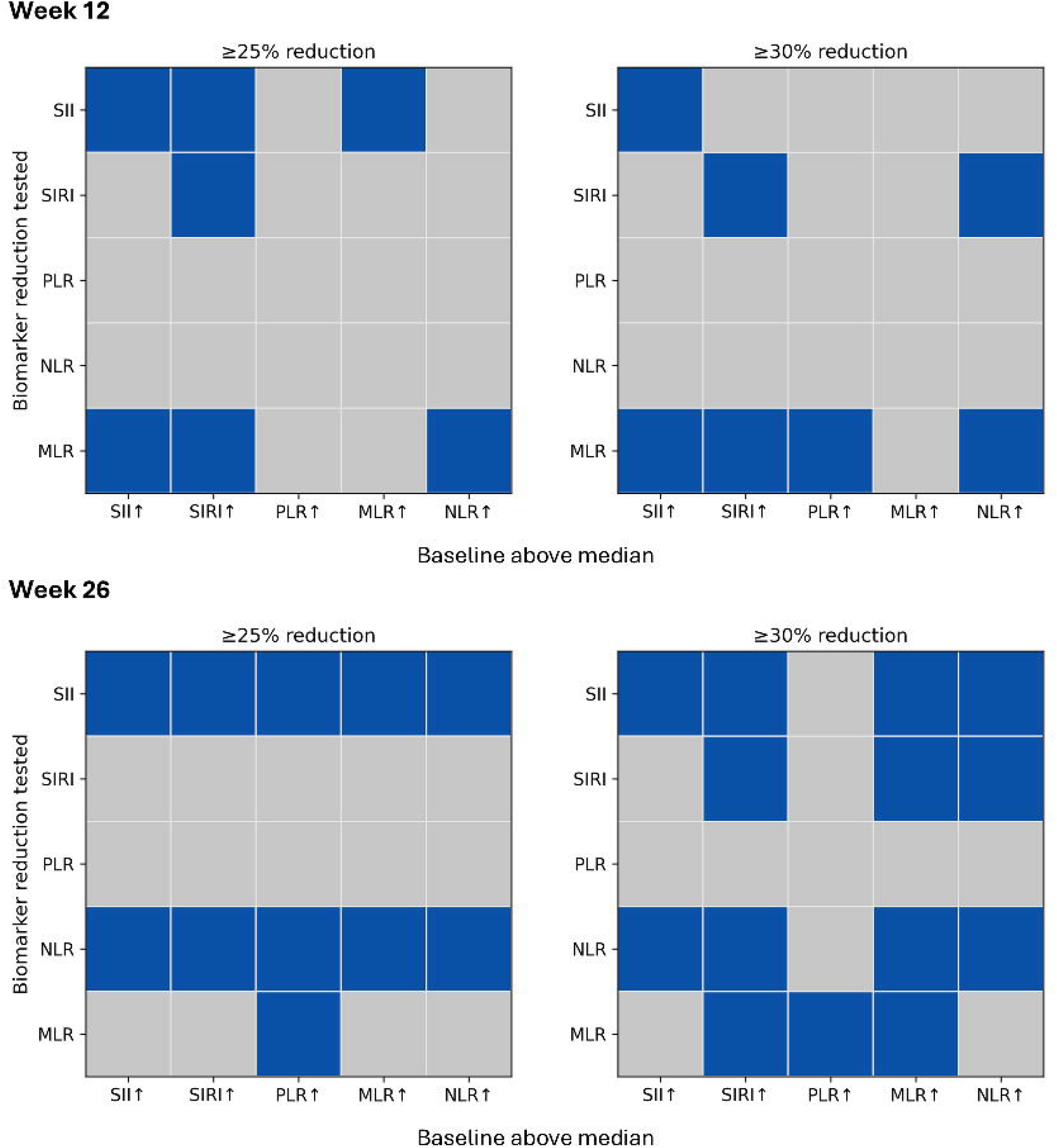
Binary matrix of statistically significant responder rates in favour of HRO over placebo for subgroups with above median baseline SII, SIRI, PLR, MLR, and NLR compared with the same parameters at week 12 and week 26. Responder criteria were defined as ≥25% or ≥30% reduction in the biomarker from baseline at the given visit. Blue tiles indicate p < 0.05 compared to placebo.

## 4 Discussion

The exploratory *post hoc* analysis of composite inflammatory biomarkers derived from CBC counts demonstrated that HRO supplementation over 26 weeks led to significant reductions in inflammation across several biomarkers compared to placebo. Notably, reductions in SII, SIRI, and MLR were observed, with evidence of an earlier onset of effect at week 12 for some markers. Sensitivity analyses using different categorical thresholds confirmed these findings, highlighting that HRO treatment resulted in meaningful reductions in inflammation, particularly for MLR at week 12.

Among the five composite inflammatory biomarkers analysed, SII was most consistently associated with reductions following HRO treatment. Stratification by baseline SII revealed that patients with low baseline values were more likely to achieve PASI50 at week 26, supporting SII as a predictive marker for HRO treatment response. This observation was aligned with other studies, as SII has been used to predict biologic therapy switching in psoriasis (22) and responsiveness to TNF-α inhibitors in rheumatoid arthritis (40) as well as predicting responsiveness to cancer immunotherapy (41). SIRI and MLR also showed significant reductions, but stratification on these markers did not correlate with improved skin symptoms, underscoring the distinct predictive value of SII.

Baseline stratification revealed that patients with higher initial values of SII, SIRI, NLR, PLR, or MLR were more likely to experience reduction in their biomarkers following HRO treatment, suggesting a modulatory effect that was most pronounced in individuals with elevated systemic inflammation at baseline. This pattern was consistent with a modulatory and inflammation-resolving effect of HRO supplementation rather than indiscriminate suppression, as the greatest treatment effects were observed in patients characterized by elevated inflammatory markers at baseline, while comparatively little change was seen in patients whose baseline values were closer to ranges associated with a more balanced immune status. The importance of this observation is highlighted by observational trials showing non-linear risk curves for SII, indicating an association between very low SII and unfavourable health outcomes due to impaired immune and platelet function (42). SII reflects systemic inflammatory load by balancing immune activation and suppression, while SIRI is more closely linked to myeloid-driven inflammation through monocyte involvement. Consistent reductions in SII and SIRI suggest a gradual cooling of the immune system, supported by *in vitro* observations of HRO’s ability to reduce IL-23 secretion (37). Reductions in MLR paralleled those observed for SIRI, consistent with their shared dependence on monocyte-driven inflammatory activity. The observed resolving effects on systemic inflammation are hypothesized to stem from HRO’s unique composition, rich in DHA and EPA in a 3:1 ratio bound in polar phospholipids, which has been associated with increased bioavailability and higher acute uptake (25). The observation that the fatty acids in HRO may contribute to reduce inflammation, is supported by data showing the omega-3 index (% omega-3 in red blood cell membranes) being negatively associated with SII (43) and dietary omega-3s negatively associated with SIRI (44).

Omega-3 fatty acids are considered to exert their inflammation-resolving effects through signalling metabolites called specialized pro-resolving mediators (SPMs) (45,46), which actively drive resolution of inflammation and restoration of tissue homeostasis. A recent publication shows that HRO contains lipid mediators and SPMs and furthermore stimulate SPM biosynthesis in immune cells *in vitro* (36). Several of the SPMs found to be biosynthesized in macrophages stimulated with HRO, including E- and D-series resolvins, are associated with reduced neutrophil recruitment and persistence (45,47–50). Consistent with this interpretation, a reduction in circulating neutrophils was also demonstrated with supplementation of HRO in a preclinical pig model (35). It should be mentioned that, in addition to effects on neutrophils, SPMs such as PDX also inhibit blood platelet aggregation, which directly would contribute to reducing SII but not SIRI (51). The statistically significant continuous-scale reductions observed in PLR supports that there also was an impact on platelet activity from the HRO treatment. Future studies should directly quantify circulating lipid mediators to establish causal links with impact on composite systemic inflammatory indices.

Although SII and SIRI are increasingly recognized as measures of systemic inflammation across several disease categories, their application has mostly been limited to observational studies, where they have been used as severity cutoffs, disease risk predictors, and as measures on probability of favourable treatment outcomes. But there have, to the authors’ knowledge, been few, if any, trials assessing the longitudinal changes of SII and SIRI in a placebo-controlled setting. Particularly not in a controlled clinical framework in patients with a confirmed IMID diagnosis. So even though the value of SII and SIRI has been confirmed as prognostic tools, there are few reports on the causal connection with a successful treatment outcome in a placebo-controlled setting. Additionally, the systemic nature of these biomarkers suggests broader health benefits, as elevated SII and SIRI has been associated with increased disease activity in various immune-mediated diseases and with risk of cardiovascular and cerebrovascular conditions (52–56). These findings support the translational relevance of HRO supplementation in conditions beyond psoriasis.

This *post hoc* analysis supports a treatment-associated effect in a placebo-controlled setting. Limitations include the exploratory nature of the analyses, lack of formal multiplicity control, and modest sample size, which may affect statistical power and confidence intervals in certain endpoints, particularly for the subgroup analyses. On the other hand, the risk of false positive findings is partly mitigated through the consistency of observed effect across different endpoints and analytical approaches. However, the findings should be regarded as hypothesis-generating and warrant further investigation in larger, confirmatory trials.

### 4.1 Conclusion

A statistically significant reduction in systemic composite inflammatory biomarkers including SII and SIRI was found in patients with mild-to-moderate psoriasis who were treated with HRO over 26 weeks compared to placebo. The reductions observed in the systemic inflammation indices were accompanied by a statistically significant clinical disease improvement as measured by mean change in PASI after 26 weeks (23).

Stratifying patients per baseline inflammatory biomarker levels was predictive of treatment response. Patients with lower baseline SII (≤518) experienced greater treatment benefit from HRO, with a higher PASI50 response rate compared to placebo. Furthermore, patients with elevated baseline inflammation (above median) experienced greater percentage-wise reduction in systemic inflammation from baseline. This may indicate that HRO works through inflammation resolution mechanisms rather than immunosuppressive processes.

The inflammatory biomarkers SII and SIRI have the potential to be used as tools for stratification in clinical trials on patients with inflammatory disorders and could be applied in future investigations of a potential role of HRO in modulating systemic inflammatory burden in other conditions with systemic inflammatory involvement.

## Data Availability

All data produced in the present study are available upon reasonable request to the authors

## 5 Abbreviations

ANCOVA: analysis of covariance
BMI: body mass index
CBC: complete blood count
CCL2: C–C motif chemokine ligand 2
CI: confidence interval
DHA: docosahexaenoic acid
DLQI: Dermatology Life Quality Index
EPA: eicosapentaenoic acid
HRO: herring roe oil
IFN□γR1: interferon gamma receptor 1
IL: interleukin
IMID: immune□mediated inflammatory disease
IQR: interquartile range
LC□PUFA: long□chain polyunsaturated fatty acid
MCT: medium□chain triglycerides
MLR: monocyte□to□lymphocyte ratio
NLR: neutrophil□to□lymphocyte ratio
OLE: open□label extension
OR: odds ratio
PASI: Psoriasis Area and Severity Index
PASI50: ≥50% reduction in PASI from baseline
PBO: placebo
PDX: protectin DX
PL: phospholipid
PLR: platelet□to□lymphocyte ratio
PUFA: polyunsaturated fatty acid
QoL: quality of life
SD: standard deviation
SE: standard error
SII: systemic immune□inflammation index
SII25/30: ≥25% and ≥30% reduction in SII from baseline, same with SIRI, NLR, PLR, MLR
SIRI: systemic inflammation response index
SPM: specialized pro□resolving mediator
TNF□α: tumor necrosis factor alpha).

## 6 Acknowledgements

We thank all patients who participated in this study. The blood samples were collected at the Research Unit for Health Surveys, Department of Clinical Science, University of Bergen, Norway.

RGammelsaeter and TRingheim-Bakka are employed by Arctic Bioscience AS who sponsored the study.

KSTveit has served as a consultant or lecturer for, or received travel support from AbbVie, Novartis, Almirall, Orion, Janssen, Mundipharma, Pfizer, Serona, Shire, Boehringer Ingelheim, Bristol Myers Squibb, Sanofi, Galderma, Leo, Medac, Arctic Bioscience, Takhzyro, and Celgene, outside the submitted work.

## 7 Author contributions

TRB wrote the original draft and was involved in data curation, formal analysis, methodology, software, and visualization. RG reviewed and edited the manuscript and was involved in conceptualization, project administration, supervision, and methodology. KST reviewed and edited the manuscript and was involved in conceptualization, investigation, methodology, resources, and supervision.

