## Supplementary material for "Resolution of systemic inflammation in psoriasis following herring roe oil treatment: a *post hoc* analysis on inflammatory biomarkers in non-severe psoriatic patients"

### 1 Supplementary Tables

Table S1: ANCOVA analysis of continuous changes in systemic inflammatory biomarkers from baseline to week 12 and week 26 in the placebo and HRO arms. Mean baseline values and mean changes (SD) are shown for SII, SIRI, NLR, PLR, and MLR. Standard errors (SE), t‑statistics, residual degrees of freedom (df), two‑sided p‑values, and 95% confidence intervals (CI) for adjusted differences are reported.

| Biomarker | Week | Baseline mean Placebo | SD | Baseline mean HRO | SD | Mean change placebo | SD | Change Mean change HRO | SD | Adj. Diff. HRO vs Placebo | SE | t | dF resid | p | CI low | CI high |
| --- | --- | --- | --- | --- | --- | --- | --- | --- | --- | --- | --- | --- | --- | --- | --- | --- |
| MLR | W12 | 0,24 | 0,08 | 0,27 | 0,13 | 0,01 | 0,12 | -0,05 | 0,11 | -0,05 | 0,03 | -1,84 | 52 | 0,0717 | -0,10 | 0,00 |
| NLR | W12 | 2,24 | 0,89 | 2,24 | 1,00 | 0,14 | 1,28 | -0,14 | 1,00 | -0,28 | 0,28 | -1,00 | 52 | 0,3217 | -0,83 | 0,28 |
| PLR | W12 | 131,22 | 36,99 | 132,13 | 44,26 | 13,34 | 38,17 | -2,73 | 28,11 | -15,92 | 9,03 | -1,76 | 52 | 0,0838 | -34,04 | 2,20 |
| SII | W12 | 563,62 | 246,35 | 569,80 | 297,89 | 49,58 | 253,90 | -42,04 | 307,45 | -88,58 | 67,26 | -1,32 | 52 | 0,1936 | -223,55 | 46,39 |
| SIRI | W12 | 1,03 | 0,51 | 1,16 | 0,79 | 0,04 | 0,67 | -0,31 | 0,82 | -0,25 | 0,15 | -1,63 | 52 | 0,1082 | -0,55 | 0,06 |
| MLR | W26 | 0,24 | 0,08 | 0,27 | 0,14 | 0,04 | 0,07 | -0,01 | 0,11 | -0,03 | 0,02 | -1,83 | 53 | 0,0734 | -0,07 | 0,00 |
| NLR | W26 | 2,24 | 0,89 | 2,31 | 1,04 | -0,10 | 0,37 | -0,30 | 0,80 | -0,18 | 0,14 | -1,30 | 53 | 0,1988 | -0,46 | 0,10 |
| PLR | W26 | 131,22 | 36,99 | 135,25 | 46,33 | 20,31 | 21,81 | 2,25 | 37,04 | -17,38 | 7,91 | -2,20 | 53 | 0,0323 | -33,24 | -1,52 |
| SII | W26 | 563,62 | 246,35 | 583,96 | 301,23 | 27,54 | 132,41 | -80,65 | 248,76 | -100,32 | 44,90 | -2,23 | 53 | 0,0297 | -190,38 | -10,27 |
| SIRI | W26 | 1,03 | 0,51 | 1,19 | 0,79 | 0,10 | 0,38 | -0,24 | 0,68 | -0,26 | 0,11 | -2,29 | 53 | 0,0263 | -0,48 | -0,03 |

Table S2: Biomarker changes from baseline to 26 weeks of treatment for the HRO group and the placebo group. Patients HRO/placebo (n): 30/30 (baseline), 26/29 (week 12), and 27/29 (week 26). Numbers are abbreviated from result spreadsheets.

|  |  | Baseline | Week 12 | | | Week 26 | | |
| --- | --- | --- | --- | --- | --- | --- | --- | --- |
| Group | Variable | Mean (SD) | Mean (SD) | ∆ | % | Mean (SD) | ∆ | % |
| HRO | SII | 623.85 (368.32) | 527.77 (235.69) | -96.09 | -15% | 503.30 (204.62) | -120.55 | -19% |
|  | SIRI | 1.23 (0.84) | 0.85 (0.38) | -0.38 | -31% | 0.95 (0.34) | -0.28 | -23% |
|  | NLR | 2.37 (1.10) | 2.11 (0.84) | -0.26 | -11% | 2.00 (0.70) | -0.36 | -15% |
|  | PLR | 137.93 (48.79) | 129.40 (42.96) | -8.53 | -6% | 137.49 (43.45) | -0.44 | 0% |
|  | MLR | 0.27 (0.13) | 0.21 (0.08) | -0,06 | -22% | 0.26 (0.08) | -0.01 | -3% |
| Placebo | SII | 561.03 (242.48) | 613.20 (306.32) | 52.17 | 9% | 591.16 (251.50) | 30.13 | 5% |
|  | SIRI | 1.03 (0.50) | 1.07 (0.75) | 0.04 | 4% | 1.13 (0.63) | 0.10 | 10% |
|  | NLR | 2.23 (0.88) | 2.38 (1.27) | 0.16 | 7% | 2.14 (0.90) | -0.09 | -4% |
|  | PLR | 132.43 (36.95) | 144.57 (48.51) | 12.13 | 9% | 151.54 (44.67) | 19.11 | 14% |
|  | MLR | 0.24 (0.09) | 0.25 (0.13) | 0.01 | 3% | 0.28 (0.09) | 0.04 | 16% |

Table S3: Categorical responder analyses for systemic inflammatory biomarkers at week 12 and week 26. Responders were defined as achieving a ≥25% or ≥30% reduction from baseline in the respective biomarker (SII, SIRI, NLR, PLR, or MLR). Odds ratios (OR) with 95% confidence intervals (CI) and two‑sided p‑values were calculated to compare responder proportions between treatment groups.

| Biomarker | Week | Threshold | n HRO | Resp. HRO | Resp. HRO % | n placebo | Resp. placebo | Resp placebo % | OR | CI 95% low | CI 95% high | p | Method |
| --- | --- | --- | --- | --- | --- | --- | --- | --- | --- | --- | --- | --- | --- |
| SII | W12 | 25% | 26 | 10 | 38% | 29 | 3 | 10% | 5,42 | 16,81 | 1,74 | 0,0245 | Fisher exact (two-sided) |
| MLR | W12 | 25% | 26 | 11 | 42% | 29 | 4 | 14% | 4,58 | 13,16 | 1,57 | 0,0321 | Fisher exact (two-sided) |
| PLR | W12 | 25% | 26 | 4 | 15% | 29 | 1 | 3% | 5,09 | 22,56 | 1,11 | 0,1777 | Fisher exact (two-sided) |
| SIRI | W12 | 25% | 26 | 11 | 42% | 29 | 8 | 28% | 1,93 | 0,62 | 5,94 | 0,2517 | Pearson chi-square |
| NLR | W12 | 25% | 26 | 8 | 31% | 29 | 7 | 24% | 1,40 | 0,42 | 4,59 | 0,5814 | Pearson chi-square |
| SII | W26 | 25% | 27 | 9 | 33% | 29 | 2 | 7% | 6,75 | 23,63 | 1,94 | 0,0184 | Fisher exact (two-sided) |
| NLR | W26 | 25% | 27 | 9 | 33% | 29 | 3 | 10% | 4,33 | 13,56 | 1,39 | 0,0519 | Fisher exact (two-sided) |
| MLR | W26 | 25% | 27 | 5 | 19% | 29 | 1 | 3% | 6,36 | 27,77 | 1,42 | 0,0959 | Fisher exact (two-sided) |
| PLR | W26 | 25% | 27 | 2 | 7% | 29 | 0 | 0% | 5,78 | 0,00 | inf | 0,2279 | Fisher exact (two-sided) |
| SIRI | W26 | 25% | 27 | 8 | 30% | 29 | 5 | 17% | 2,02 | 5,69 | 0,71 | 0,3490 | Fisher exact (two-sided) |
| MLR | W12 | 30% | 26 | 10 | 38% | 29 | 2 | 7% | 8,44 | 29,30 | 2,43 | 0,0076 | Fisher exact (two-sided) |
| MLR | W26 | 30% | 27 | 5 | 19% | 29 | 0 | 0% | 14,42 | 0,00 | inf | 0,0211 | Fisher exact (two-sided) |
| NLR | W26 | 30% | 27 | 7 | 26% | 29 | 1 | 3% | 9,80 | 41,87 | 2,23 | 0,0227 | Fisher exact (two-sided) |
| SII | W26 | 30% | 27 | 7 | 26% | 29 | 1 | 3% | 9,80 | 41,87 | 2,23 | 0,0227 | Fisher exact (two-sided) |
| SIRI | W12 | 30% | 26 | 10 | 38% | 29 | 3 | 10% | 5,42 | 16,81 | 1,74 | 0,0245 | Fisher exact (two-sided) |
| SII | W12 | 30% | 26 | 8 | 31% | 29 | 3 | 10% | 3,85 | 12,18 | 1,22 | 0,0915 | Fisher exact (two-sided) |
| SIRI | W26 | 30% | 27 | 7 | 26% | 29 | 3 | 10% | 3,03 | 9,71 | 0,95 | 0,1710 | Fisher exact (two-sided) |
| PLR | W26 | 30% | 27 | 2 | 7% | 29 | 0 | 0% | 5,78 | 0,00 | inf | 0,2279 | Fisher exact (two-sided) |
| NLR | W12 | 30% | 26 | 6 | 23% | 29 | 3 | 10% | 2,60 | 8,47 | 0,80 | 0,2808 | Fisher exact (two-sided) |
| PLR | W12 | 30% | 26 | 1 | 4% | 29 | 0 | 0% | 3,47 | 0,00 | inf | 0,4727 | Fisher exact (two-sided) |

Table S4: Subgroup analyses of clinical responder endpoints stratified by baseline systemic inflammatory biomarkers. PASI50 (≥50% reduction in PASI from baseline) and clinically meaningful improvement in quality of life (∆DLQI ≤ −4) were evaluated at week 12 and week 26 in subgroups defined by biomarker values at or below the population median for SII, SIRI, NLR, PLR, or MLR. Odds ratios (OR) with 95% confidence intervals (CI) and two‑sided p‑values were calculated to compare responder proportions between treatment groups.

| Endpoint | Week | Subgroup | n HRO | Resp. HRO | Resp. HRO % | n placebo | Resp. placebo | Resp. Placebo % | OR | CI 95% low | CI 95% high | p |
| --- | --- | --- | --- | --- | --- | --- | --- | --- | --- | --- | --- | --- |
| PASI50 | W26 | ≤ median SII | 16 | 7 | 44% | 13 | 1 | 8% | 9,33 | 39,23 | 1,99 | 0,0443 |
| PASI50 | W26 | ≤ median NLR | 13 | 6 | 46% | 16 | 2 | 13% | 6,00 | 21,70 | 1,52 | 0,0923 |
| PASI50 | W26 | ≤ median MLR | 14 | 6 | 43% | 14 | 2 | 14% | 4,50 | 16,46 | 1,15 | 0,2087 |
| PASI50 | W26 | ≤ median SIRI | 14 | 6 | 43% | 15 | 3 | 20% | 3,00 | 10,13 | 0,85 | 0,2451 |
| PASI50 | W26 | ≤ median PLR | 14 | 4 | 29% | 14 | 1 | 7% | 5,20 | 22,70 | 1,09 | 0,3259 |
| PASI50 | W12 | ≤ median SII | 16 | 2 | 13% | 13 | 0 | 0% | 4,66 | 0,00 | inf | 0,4877 |
| PASI50 | W12 | ≤ median SIRI | 14 | 1 | 7% | 15 | 1 | 7% | 1,08 | 5,65 | 0,20 | 1,0000 |
| PASI50 | W12 | ≤ median NLR | 13 | 1 | 8% | 16 | 1 | 6% | 1,25 | 6,53 | 0,24 | 1,0000 |
| PASI50 | W12 | ≤ median PLR | 15 | 0 | 0% | 14 | 0 | 0% | 0,94 | 0,00 | inf | 1,0000 |
| PASI50 | W12 | ≤ median MLR | 15 | 1 | 7% | 14 | 1 | 7% | 0,93 | 4,90 | 0,18 | 1,0000 |
| ∆DLQI ≤-4 | W12 | ≤ median MLR | 15 | 4 | 27% | 14 | 7 | 50% | 0,36 | 1,23 | 0,11 | 0,2635 |
| ∆DLQI ≤-4 | W12 | ≤ median SIRI | 14 | 3 | 21% | 15 | 6 | 40% | 0,41 | 1,44 | 0,12 | 0,4270 |
| ∆DLQI ≤-4 | W12 | ≤ median NLR | 13 | 3 | 23% | 16 | 7 | 44% | 0,39 | 1,34 | 0,11 | 0,4335 |
| ∆DLQI ≤-4 | W26 | ≤ median NLR | 13 | 3 | 23% | 16 | 7 | 44% | 0,39 | 1,34 | 0,11 | 0,4335 |
| ∆DLQI ≤-4 | W26 | ≤ median MLR | 14 | 4 | 29% | 14 | 6 | 43% | 0,53 | 1,80 | 0,16 | 0,6946 |
| ∆DLQI ≤-4 | W12 | ≤ median SII | 16 | 3 | 19% | 13 | 3 | 23% | 0,77 | 2,91 | 0,21 | 1,0000 |
| ∆DLQI ≤-4 | W12 | ≤ median PLR | 15 | 4 | 27% | 14 | 4 | 29% | 0,91 | 3,13 | 0,27 | 1,0000 |
| ∆DLQI ≤-4 | W26 | ≤ median SII | 16 | 4 | 25% | 13 | 3 | 23% | 1,11 | 4,01 | 0,31 | 1,0000 |
| ∆DLQI ≤-4 | W26 | ≤ median SIRI | 14 | 4 | 29% | 15 | 5 | 33% | 0,80 | 2,69 | 0,24 | 1,0000 |
| ∆DLQI ≤-4 | W26 | ≤ median PLR | 14 | 4 | 29% | 14 | 4 | 29% | 1,00 | 3,45 | 0,29 | 1,0000 |

Table S5: Cross‑stratified categorical responder analyses for systemic inflammatory biomarkers at week 12 and week 26. Responders were defined as achieving a ≥25% or ≥30% reduction from baseline in the indicated biomarker, within subgroups stratified by baseline values, defined relative to the population median. Odds ratios (OR) with 95% confidence intervals (CI) and two‑sided p‑values compare responder proportions between treatment groups.

| Biomarker | Week | Threshold | Biomarker strata | n HRO | Resp. HRO | Resp. HRO % | n placebo | Resp. placebo | Resp. placebo % | OR | CI 95% low | CI 95% high | p |
| --- | --- | --- | --- | --- | --- | --- | --- | --- | --- | --- | --- | --- | --- |
| MLR | W12 | 25% | SIRI | 12 | 8 | 67 | 14 | 2 | 14 | 12,00 | 41,89 | 2,83 | 0,0138 |
| SII | W12 | 25% | MLR | 12 | 6 | 50 | 15 | 1 | 7 | 14,00 | 57,35 | 2,84 | 0,0237 |
| SII | W12 | 25% | SIRI | 12 | 6 | 50 | 14 | 1 | 7 | 13,00 | 53,36 | 2,63 | 0,0261 |
| SII | W12 | 25% | SII | 10 | 6 | 60 | 17 | 3 | 18 | 7,00 | 23,61 | 1,79 | 0,0393 |
| SIRI | W12 | 25% | SIRI | 12 | 8 | 67 | 14 | 3 | 21 | 7,33 | 24,41 | 1,90 | 0,0447 |
| MLR | W12 | 25% | NLR | 13 | 7 | 54 | 14 | 2 | 14 | 7,00 | 25,05 | 1,75 | 0,0461 |
| MLR | W12 | 25% | SII | 10 | 4 | 40 | 17 | 1 | 6 | 10,67 | 44,72 | 2,12 | 0,0473 |
| SII | W12 | 25% | PLR | 12 | 7 | 58 | 15 | 3 | 20 | 5,60 | 18,68 | 1,50 | 0,0568 |
| SII | W12 | 25% | NLR | 13 | 6 | 46 | 14 | 2 | 14 | 5,14 | 18,74 | 1,30 | 0,1032 |
| MLR | W12 | 25% | PLR | 12 | 6 | 50 | 15 | 3 | 20 | 4,00 | 13,53 | 1,09 | 0,1266 |
| NLR | W12 | 25% | PLR | 12 | 6 | 50 | 15 | 3 | 20 | 4,00 | 13,53 | 1,09 | 0,1266 |
| MLR | W12 | 25% | MLR | 12 | 7 | 58 | 15 | 4 | 27 | 3,85 | 12,49 | 1,09 | 0,1302 |
| PLR | W12 | 25% | PLR | 12 | 4 | 33 | 15 | 1 | 7 | 7,00 | 30,09 | 1,44 | 0,1390 |
| NLR | W12 | 25% | SIRI | 12 | 6 | 50 | 14 | 3 | 21 | 3,67 | 12,47 | 1,00 | 0,2177 |
| NLR | W12 | 25% | SII | 10 | 6 | 60 | 17 | 5 | 29 | 3,60 | 11,81 | 1,01 | 0,2238 |
| SIRI | W12 | 25% | MLR | 12 | 7 | 58 | 15 | 5 | 33 | 2,80 | 8,98 | 0,82 | 0,2576 |
| NLR | W12 | 25% | MLR | 12 | 5 | 42 | 15 | 3 | 20 | 2,86 | 9,87 | 0,78 | 0,3981 |
| NLR | W12 | 25% | NLR | 13 | 6 | 46 | 14 | 4 | 29 | 2,14 | 7,03 | 0,63 | 0,4401 |
| PLR | W12 | 25% | SII | 10 | 2 | 20 | 17 | 1 | 6 | 4,00 | 18,36 | 0,78 | 0,5350 |
| SIRI | W12 | 25% | NLR | 13 | 7 | 54 | 14 | 6 | 43 | 1,56 | 0,34 | 7,11 | 0,5680 |
| PLR | W12 | 25% | MLR | 12 | 2 | 17 | 15 | 1 | 7 | 2,80 | 13,07 | 0,55 | 0,5692 |
| PLR | W12 | 25% | SIRI | 12 | 2 | 17 | 14 | 1 | 7 | 2,60 | 12,16 | 0,51 | 0,5800 |
| PLR | W12 | 25% | NLR | 13 | 2 | 15 | 14 | 1 | 7 | 2,36 | 11,11 | 0,47 | 0,5956 |
| SIRI | W12 | 25% | SII | 10 | 5 | 50 | 17 | 6 | 35 | 1,83 | 6,01 | 0,54 | 0,6868 |
| SIRI | W12 | 25% | PLR | 12 | 6 | 50 | 15 | 7 | 47 | 1,14 | 0,25 | 5,22 | 0,8632 |
| SII | W26 | 25% | NLR | 14 | 9 | 64 | 14 | 0 | 0 | 50,09 | 0,00 | inf | 0,0006 |
| SII | W26 | 25% | SII | 11 | 7 | 64 | 17 | 1 | 6 | 28,00 | 108,85 | 5,38 | 0,0019 |
| SII | W26 | 25% | SIRI | 13 | 7 | 54 | 14 | 0 | 0 | 33,46 | 0,00 | inf | 0,0019 |
| SII | W26 | 25% | MLR | 13 | 6 | 46 | 15 | 0 | 0 | 26,87 | 0,00 | inf | 0,0046 |
| SII | W26 | 25% | PLR | 13 | 7 | 54 | 15 | 1 | 7 | 16,33 | 66,38 | 3,33 | 0,0108 |
| NLR | W26 | 25% | NLR | 14 | 8 | 57 | 14 | 1 | 7 | 17,33 | 70,16 | 3,55 | 0,0128 |
| NLR | W26 | 25% | SIRI | 13 | 7 | 54 | 14 | 1 | 7 | 15,17 | 61,76 | 3,09 | 0,0128 |
| NLR | W26 | 25% | PLR | 13 | 6 | 46 | 15 | 1 | 7 | 12,00 | 49,77 | 2,48 | 0,0286 |
| NLR | W26 | 25% | MLR | 13 | 6 | 46 | 15 | 1 | 7 | 12,00 | 49,77 | 2,48 | 0,0286 |
| NLR | W26 | 25% | SII | 11 | 6 | 55 | 17 | 2 | 12 | 9,00 | 32,09 | 2,18 | 0,0299 |
| MLR | W26 | 25% | PLR | 13 | 4 | 31 | 15 | 0 | 0 | 14,68 | 0,00 | inf | 0,0349 |
| MLR | W26 | 25% | SII | 11 | 3 | 27 | 17 | 0 | 0 | 14,41 | 0,00 | inf | 0,0504 |
| MLR | W26 | 25% | NLR | 14 | 4 | 29 | 14 | 0 | 0 | 12,43 | 0,00 | inf | 0,0978 |
| SIRI | W26 | 25% | SIRI | 13 | 7 | 54 | 14 | 3 | 21 | 4,28 | 14,32 | 1,17 | 0,1201 |
| SIRI | W26 | 25% | NLR | 14 | 8 | 57 | 14 | 3 | 21 | 4,89 | 16,17 | 1,35 | 0,1201 |
| MLR | W26 | 25% | MLR | 13 | 4 | 31 | 15 | 1 | 7 | 6,22 | 26,95 | 1,29 | 0,1528 |
| MLR | W26 | 25% | SIRI | 13 | 4 | 31 | 14 | 1 | 7 | 5,78 | 25,07 | 1,20 | 0,1647 |
| SIRI | W26 | 25% | MLR | 13 | 7 | 54 | 15 | 4 | 27 | 3,21 | 10,32 | 0,93 | 0,2458 |
| SIRI | W26 | 25% | SII | 11 | 5 | 45 | 17 | 4 | 24 | 2,71 | 8,98 | 0,77 | 0,4087 |
| SIRI | W26 | 25% | PLR | 13 | 6 | 46 | 15 | 4 | 27 | 2,36 | 7,68 | 0,69 | 0,4328 |
| PLR | W26 | 25% | PLR | 13 | 1 | 8 | 15 | 0 | 0 | 3,72 | 0,00 | inf | 0,4643 |
| PLR | W26 | 25% | MLR | 13 | 1 | 8 | 15 | 0 | 0 | 3,72 | 0,00 | inf | 0,4643 |
| PLR | W26 | 25% | SIRI | 13 | 1 | 8 | 14 | 0 | 0 | 3,48 | 0,00 | inf | 0,4815 |
| PLR | W26 | 25% | NLR | 14 | 2 | 14 | 14 | 0 | 0 | 5,80 | 0,00 | inf | 0,4815 |
| PLR | W26 | 25% | SII | 11 | 0 | 0 | 17 | 0 | 0 | 1,52 | 0,00 | inf | 1,0000 |
| SIRI | W12 | 30% | SIRI | 12 | 8 | 67 | 14 | 1 | 7 | 26,00 | 100,99 | 5,00 | 0,0029 |
| MLR | W12 | 30% | SIRI | 12 | 7 | 58 | 14 | 1 | 7 | 18,20 | 72,88 | 3,62 | 0,0093 |
| MLR | W12 | 30% | PLR | 12 | 6 | 50 | 15 | 1 | 7 | 14,00 | 57,35 | 2,84 | 0,0237 |
| MLR | W12 | 30% | NLR | 13 | 6 | 46 | 14 | 1 | 7 | 11,14 | 46,31 | 2,30 | 0,0329 |
| SII | W12 | 30% | SII | 10 | 6 | 60 | 17 | 3 | 18 | 7,00 | 23,61 | 1,79 | 0,0393 |
| SIRI | W12 | 30% | NLR | 13 | 7 | 54 | 14 | 2 | 14 | 7,00 | 25,05 | 1,75 | 0,0461 |
| MLR | W12 | 30% | SII | 10 | 4 | 40 | 17 | 1 | 6 | 10,67 | 44,72 | 2,12 | 0,0473 |
| SIRI | W12 | 30% | SII | 10 | 5 | 50 | 17 | 2 | 12 | 7,50 | 27,21 | 1,79 | 0,0646 |
| NLR | W12 | 30% | SIRI | 12 | 5 | 42 | 14 | 1 | 7 | 9,29 | 39,01 | 1,90 | 0,0652 |
| NLR | W12 | 30% | NLR | 13 | 5 | 38 | 14 | 1 | 7 | 8,13 | 34,46 | 1,69 | 0,0768 |
| MLR | W12 | 30% | MLR | 12 | 6 | 50 | 15 | 2 | 13 | 6,50 | 23,47 | 1,61 | 0,0870 |
| SIRI | W12 | 30% | PLR | 12 | 6 | 50 | 15 | 2 | 13 | 6,50 | 23,47 | 1,61 | 0,0870 |
| SIRI | W12 | 30% | MLR | 12 | 6 | 50 | 15 | 2 | 13 | 6,50 | 23,47 | 1,61 | 0,0870 |
| SII | W12 | 30% | PLR | 12 | 6 | 50 | 15 | 3 | 20 | 4,00 | 13,53 | 1,09 | 0,1266 |
| NLR | W12 | 30% | PLR | 12 | 4 | 33 | 15 | 1 | 7 | 7,00 | 30,09 | 1,44 | 0,1390 |
| NLR | W12 | 30% | MLR | 12 | 4 | 33 | 15 | 1 | 7 | 7,00 | 30,09 | 1,44 | 0,1390 |
| SII | W12 | 30% | MLR | 12 | 4 | 33 | 15 | 1 | 7 | 7,00 | 30,09 | 1,44 | 0,1390 |
| SII | W12 | 30% | SIRI | 12 | 4 | 33 | 14 | 1 | 7 | 6,50 | 28,00 | 1,33 | 0,1478 |
| NLR | W12 | 30% | SII | 10 | 4 | 40 | 17 | 2 | 12 | 5,00 | 18,72 | 1,20 | 0,1535 |
| SII | W12 | 30% | NLR | 13 | 5 | 38 | 14 | 2 | 14 | 3,75 | 13,96 | 0,95 | 0,2087 |
| PLR | W12 | 30% | SII | 10 | 1 | 10 | 17 | 0 | 0 | 5,53 | 0,00 | inf | 0,3704 |
| PLR | W12 | 30% | PLR | 12 | 1 | 8 | 15 | 0 | 0 | 4,04 | 0,00 | inf | 0,4444 |
| PLR | W12 | 30% | NLR | 13 | 1 | 8 | 14 | 0 | 0 | 3,48 | 0,00 | inf | 0,4815 |
| PLR | W12 | 30% | SIRI | 12 | 0 | 0 | 14 | 0 | 0 | 1,16 | 0,00 | inf | 1,0000 |
| PLR | W12 | 30% | MLR | 12 | 0 | 0 | 15 | 0 | 0 | 1,24 | 0,00 | inf | 1,0000 |
| NLR | W26 | 30% | SIRI | 13 | 6 | 46 | 14 | 0 | 0 | 25,13 | 0,00 | inf | 0,0058 |
| NLR | W26 | 30% | NLR | 14 | 7 | 50 | 14 | 0 | 0 | 29,00 | 0,00 | inf | 0,0058 |
| SII | W26 | 30% | NLR | 14 | 7 | 50 | 14 | 0 | 0 | 29,00 | 0,00 | inf | 0,0058 |
| NLR | W26 | 30% | MLR | 13 | 5 | 38 | 15 | 0 | 0 | 20,06 | 0,00 | inf | 0,0131 |
| SII | W26 | 30% | SIRI | 13 | 5 | 38 | 14 | 0 | 0 | 18,76 | 0,00 | inf | 0,0159 |
| NLR | W26 | 30% | SII | 11 | 5 | 45 | 17 | 1 | 6 | 13,33 | 55,03 | 2,68 | 0,0221 |
| SII | W26 | 30% | SII | 11 | 5 | 45 | 17 | 1 | 6 | 13,33 | 55,03 | 2,68 | 0,0221 |
| SIRI | W26 | 30% | NLR | 14 | 7 | 50 | 14 | 1 | 7 | 13,00 | 53,60 | 2,70 | 0,0329 |
| MLR | W26 | 30% | PLR | 13 | 4 | 31 | 15 | 0 | 0 | 14,68 | 0,00 | inf | 0,0349 |
| MLR | W26 | 30% | MLR | 13 | 4 | 31 | 15 | 0 | 0 | 14,68 | 0,00 | inf | 0,0349 |
| SII | W26 | 30% | MLR | 13 | 4 | 31 | 15 | 0 | 0 | 14,68 | 0,00 | inf | 0,0349 |
| MLR | W26 | 30% | SIRI | 13 | 4 | 31 | 14 | 0 | 0 | 13,74 | 0,00 | inf | 0,0407 |
| SIRI | W26 | 30% | MLR | 13 | 7 | 54 | 15 | 2 | 13 | 7,58 | 27,03 | 1,90 | 0,0418 |
| SIRI | W26 | 30% | SIRI | 13 | 7 | 54 | 14 | 2 | 14 | 7,00 | 25,05 | 1,75 | 0,0461 |
| MLR | W26 | 30% | SII | 11 | 3 | 27 | 17 | 0 | 0 | 14,41 | 0,00 | inf | 0,0504 |
| SII | W26 | 30% | PLR | 13 | 5 | 38 | 15 | 1 | 7 | 8,75 | 37,03 | 1,82 | 0,0691 |
| SIRI | W26 | 30% | SII | 11 | 5 | 45 | 17 | 2 | 12 | 6,25 | 22,82 | 1,53 | 0,0764 |
| SIRI | W26 | 30% | PLR | 13 | 6 | 46 | 15 | 2 | 13 | 5,57 | 20,22 | 1,41 | 0,0957 |
| MLR | W26 | 30% | NLR | 14 | 4 | 29 | 14 | 0 | 0 | 12,43 | 0,00 | inf | 0,0978 |
| NLR | W26 | 30% | PLR | 13 | 4 | 31 | 15 | 1 | 7 | 6,22 | 26,95 | 1,29 | 0,1528 |
| PLR | W26 | 30% | PLR | 13 | 1 | 8 | 15 | 0 | 0 | 3,72 | 0,00 | inf | 0,4643 |
| PLR | W26 | 30% | MLR | 13 | 1 | 8 | 15 | 0 | 0 | 3,72 | 0,00 | inf | 0,4643 |
| PLR | W26 | 30% | SIRI | 13 | 1 | 8 | 14 | 0 | 0 | 3,48 | 0,00 | inf | 0,4815 |
| PLR | W26 | 30% | NLR | 14 | 2 | 14 | 14 | 0 | 0 | 5,80 | 0,00 | inf | 0,4815 |
| PLR | W26 | 30% | SII | 11 | 0 | 0 | 17 | 0 | 0 | 1,52 | 0,00 | inf | 1,0000 |
